# The effectiveness of tests to detect the presence of SARS-CoV-2 virus, and antibodies to SARS-CoV-2, to inform COVID-19 diagnosis: a rapid systematic review

**DOI:** 10.1101/2020.08.10.20171777

**Authors:** David Jarrom, Lauren Elston, Jennifer Washington, Matthew Prettyjohns, Kimberley Cann, Susan Myles, Peter Groves

## Abstract

**Objectives:** We undertook a rapid systematic review with the aim of identifying evidence that could be used to answer the following research questions: (1) What is the clinical effectiveness of tests that detect the presence of severe acute respiratory syndrome coronavirus 2 (SARS-CoV-2) to inform COVID-19 diagnosis? (2) What is the clinical effectiveness of tests that detect the presence of antibodies to the SARS-CoV-2 virus to inform COVID-19 diagnosis?

**Design:** systematic review and meta-analysis of studies of diagnostic test accuracy. We systematically searched for all published evidence on the effectiveness of tests for the presence of SARS-Cov-2 virus, or antibodies to SARS-CoV-2, up to 4 May 2020, and assessed relevant studies for risks of bias using the QUADAS-2 framework.

**Main outcome measures:** measures of diagnostic accuracy (sensitivity, specificity, positive/negative predictive value) were the main outcomes of interest. We also included studies that reported influence of testing on subsequent patient management, and that reported virus/antibody detection rates where these facilitated comparisons of testing in different settings, different populations, or using different sampling methods.

**Results:** 38 studies on SARS-CoV-2 virus testing and 25 studies on SARS-CoV-2 antibody testing were identified. We identified high or unclear risks of bias in the majority of studies, most commonly as a result of unclear methods of patient selection and test conduct, or because of the use of a reference standard that may not definitively diagnose COVID-19. The majority were in hospital settings, in patients with confirmed or suspected COVID-19 infection. Pooled analysis of 16 studies (3818 patients) estimated a sensitivity of 87.8% (95% confidence interval 81.5% to 92.2%) for an initial reverse-transcriptase polymerase chain reaction test. For antibody tests, ten studies reported diagnostic accuracy outcomes: sensitivity ranged from 18.4% to 96.1% and specificity 88.9% to 100%. However, the lack of a true reference standard for SARS-CoV-2 diagnosis makes it challenging to assess the true diagnostic accuracy of these tests. Eighteen studies reporting different sampling methods suggest that for virus tests, the type of sample obtained/type of tissue sampled could influence test accuracy. Finally we searched for, but did not identify, any evidence on how any test influences subsequent patient management.

**Conclusions:** Evidence is rapidly emerging on the effectiveness of tests for COVID-19 diagnosis and management, but important uncertainties about their effectiveness and most appropriate application remain. Estimates of diagnostic accuracy should be interpreted bearing in mind the absence of a definitive reference standard to diagnose or rule out COVID-19 infection. More evidence is needed about the effectiveness of testing outside of hospital settings and in mild or asymptomatic cases. Implementation of public health strategies centred on COVID-19 testing provides opportunities to explore these important areas of research.

**SUMMARY BOX:** *What is already known about this subject?:*

- Tests for the presence of the SARS-CoV-2 virus, and antibodies to the virus, are being deployed rapidly and at scale as part of the global response to COVID-19.
- At the outset of this work (March 2020), no high-quality evidence reviews on the effectiveness of SARS-CoV-2 virus or antibody tests were available.
- High-quality evidence reviews are required to help decision makers deploy and interpret these tests effectively.

*What are the new findings?:*

- Here, we synthesise evidence on the diagnostic accuracy of all known tests for SARS-CoV-2, as well as tests for antibodies to SARS-CoV-2.
- We also systematically summarise evidence on the influence of tissue sample site on virus test detection rates, and the influence of test timing relative to disease course on antibody detection. The results suggest that both these factors could influence test results.
- We conclude that evidence on SARS-CoV-2 virus and antibody tests is nascent and significant uncertainties remain in the evidence base regarding their clinical and public health application. We also note that potential risks of bias exist within many of the available studies.

*How might it impact on clinical practice in the foreseeable future?:*

- In a rapidly developing pandemic, the widespread use of testing is an essential element in the development of effective public health strategies, but it is important to acknowledge the gaps and limitations that exist in the current evidence base and that, where possible, these should be addressed in future studies.
- In particular, more evidence is needed on the performance of point-of-care or near-patient tests compared to their laboratory equivalents, and results of testing in people with no or minimal symptoms in community-based settings needs further analysis.

## INTRODUCTION

In December 2019, a novel coronavirus was discovered in Wuhan, China, which has since spread rapidly across the world. This virus was named severe acute respiratory syndrome coronavirus 2 (SARS-CoV-2), and the disease that it causes, COVID-19. Early on in the pandemic, the World Health Organization (WHO) stated that testing for the virus should be considered for symptomatic patients on the basis of the suspicion and likelihood of COVID-19, as well as in those who are asymptomatic or minimally symptomatic but who have been in contact with confirmed cases.[1] More recently, WHO highlighted the importance of testing for disease surveillance, to limit the spread of the disease and to manage COVID-19 risk during attempts to restore normal economic and social functioning.[2] Furthermore, the Organization for Economic Co-operation and Development have identified the potential importance of testing when combined with effective contact tracing in suppressing local outbreaks of COVID-19 as well as in determining individuals who have been previously infected who may safely re-integrate into work and healthcare environments.[3]

Tests for COVID-19 fall into two broad groups: tests that detect the presence of SARS-CoV-2 virus and tests that detect the presence of antibodies to SARS-CoV-2. Tests for the presence of virus usually use methods that recognise and amplify SARS-CoV-2 viral nucleic acid, such as reverse-transcriptase polymerase chain reaction (RT-PCR) or isothermal amplification. SARS CoV-2 virus testing is usually done in a specialised laboratory setting using respiratory samples, such as nasopharyngeal swabs, but near-patient tests have also been developed. SARS CoV-2 antibody testing (also called serology testing) is done on blood or serum samples and tests have been developed both for analysis in a laboratory and a near-patient setting. Since antibodies are produced as part of the body’s immune response to infection, serology tests may be useful to identify ongoing, recovering (convalescent) or previous SARS-CoV-2 infection.

The validation and application of the different tests for COVID-19, whether for individual clinical decision-making or population-based public health strategies, is dependent on the accuracy and performance of these tests. The purpose of this review is to identify, appraise and summarise the published evidence on the diagnostic performance and effectiveness of SARS CoV-2 virus and antibody tests in the diagnosis and management of current or previous COVID-19. The review also explores the influence of a range of factors on test outcomes, such as the timing of testing relative to first diagnosis/symptom onset, sampling methods, and whether testing is laboratory-based or done at point-of-care.

## METHODS

We systematically searched for evidence to answer the following questions:

1. What is the clinical effectiveness of tests that detect the presence of the SARS-CoV-2 virus to inform COVID-19 diagnosis?
2. What is the clinical effectiveness of tests that detect the presence of antibodies to the SARS-CoV-2 virus to inform COVID-19 diagnosis?

Searching and screening for both questions was undertaken based on one search strategy. Initial scoping-level evidence searches were conducted using online databases set up to aggregate COVID-19-specific evidence.[4-6]

Based on the results of these, a specific search strategy (Supplementary Appendix 1; developed and run by JW) was used to capture published evidence on SARS-CoV-2 diagnostics. The databases searched were Medline, Embase, Cochrane Library, International Network of Agencies for Health Technology Assessment (INAHTA)HTA database & Open Grey, to include all evidence published up to 4 May 2020. The sources included in the Health Technology Wales COVID-19 Evidence Digest[7] were hand-searched for relevant evidence and key stakeholders in Wales contacted for any published or unpublished data of relevance to this review. Because this was a rapid review, the protocol was not prospectively published.

Articles were included that studied any test to detect the presence of SARS-CoV-2, or antibodies to SARS-CoV-2, in people suspected of having recent or ongoing infection, and reported detection rates, influence of test result on changes in patient management, or diagnostic accuracy. For the latter outcome, we included studies that used any suitable reference standard method of diagnosis (we excluded studies that used CT scan results alone as a reference standard). The detailed criteria used to select evidence are provided in Supplementary Appendix 1. The following data was extracted from all studies deemed relevant: study design; number of centres and their location(s); dates of enrolment; inclusion/exclusion criteria; number of patients included; age and sex of included patients; test type; test target; test supplier or manufacturer; reference standard; and outcome data for each relevant outcome reported. We used the QUADAS-2 tool to assess risk of bias and applicability of relevant articles.[8] Two authors (DJ and LE) screened studies, extracted data and carried out QUADAS-2 assessments; results were checked by a third author (KC) and any disagreements were resolved by consensus.

Meta-analysis of diagnostic accuracy outcomes (sensitivity and/or specificity) was conducted only for suitable studies that reported numbers of true and false positive and negative results validated against a suitable reference standard. Pooled estimates were calculated for diagnostic accuracy outcomes using a random effects bivariate binomial model in MetaDTA v1.25.[9]

## RESULTS

Figure 1 summarises articles included and excluded at each stage, and reasons why studies were excluded. A total of 13,677 unique articles were screened for eligibility, of which 13,285 were excluded after reading the title and abstract because they did not meet our inclusion criteria. The full text of the remaining 392 articles were read and checked for eligibility, and a further 329 were excluded. Of the remaining 63 relevant articles, 38 studied virus tests and 25 studied antibody tests. Tables 1 and 2 summarise the design and characteristics of studies reporting diagnostic accuracy outcomes for virus and antibody tests, respectively. Characteristics of studies that reported other outcomes are reported in Supplementary Appendix 2.

**Figure 1.**
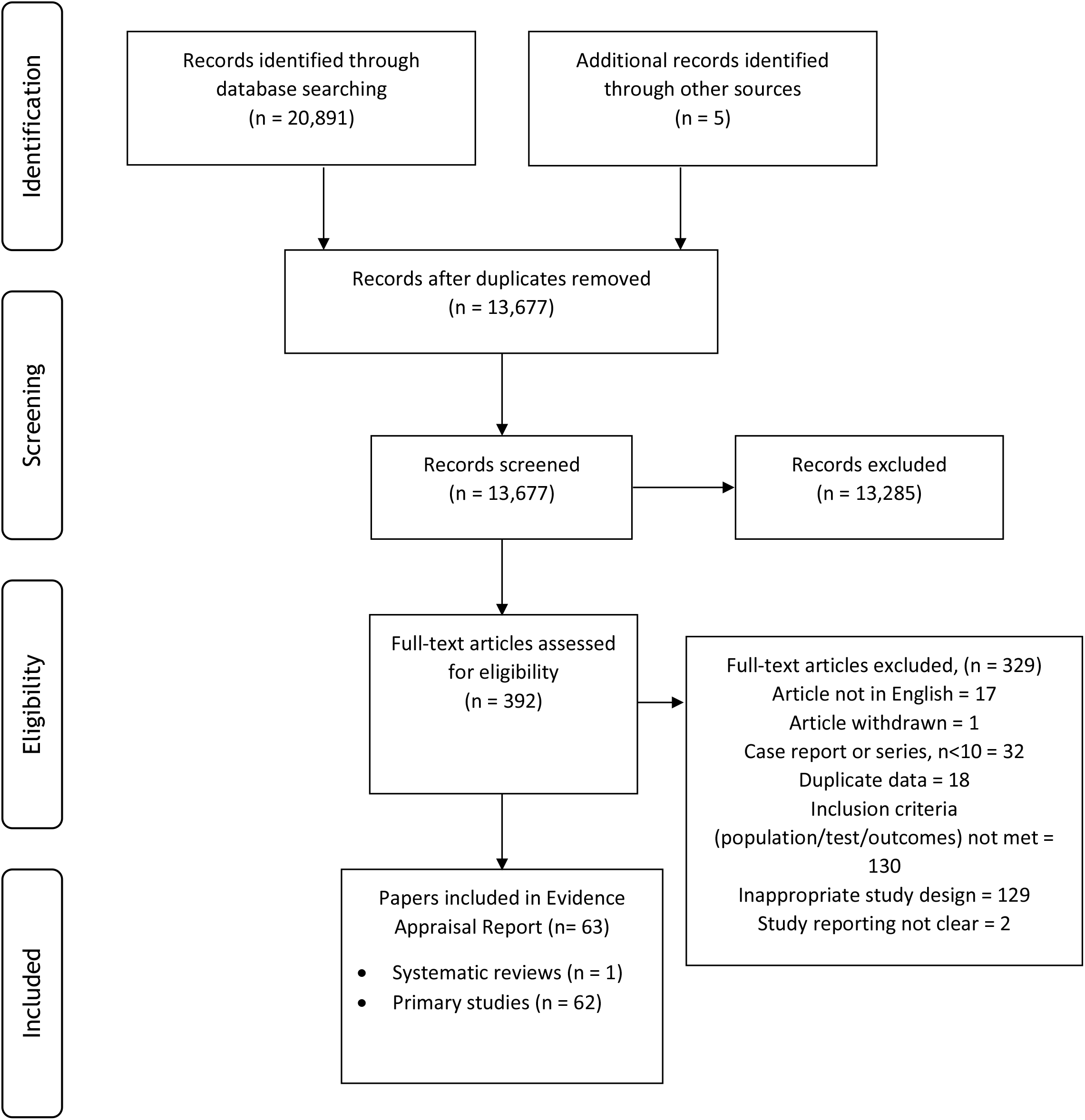
Summary of study selection.

**Table 1.**
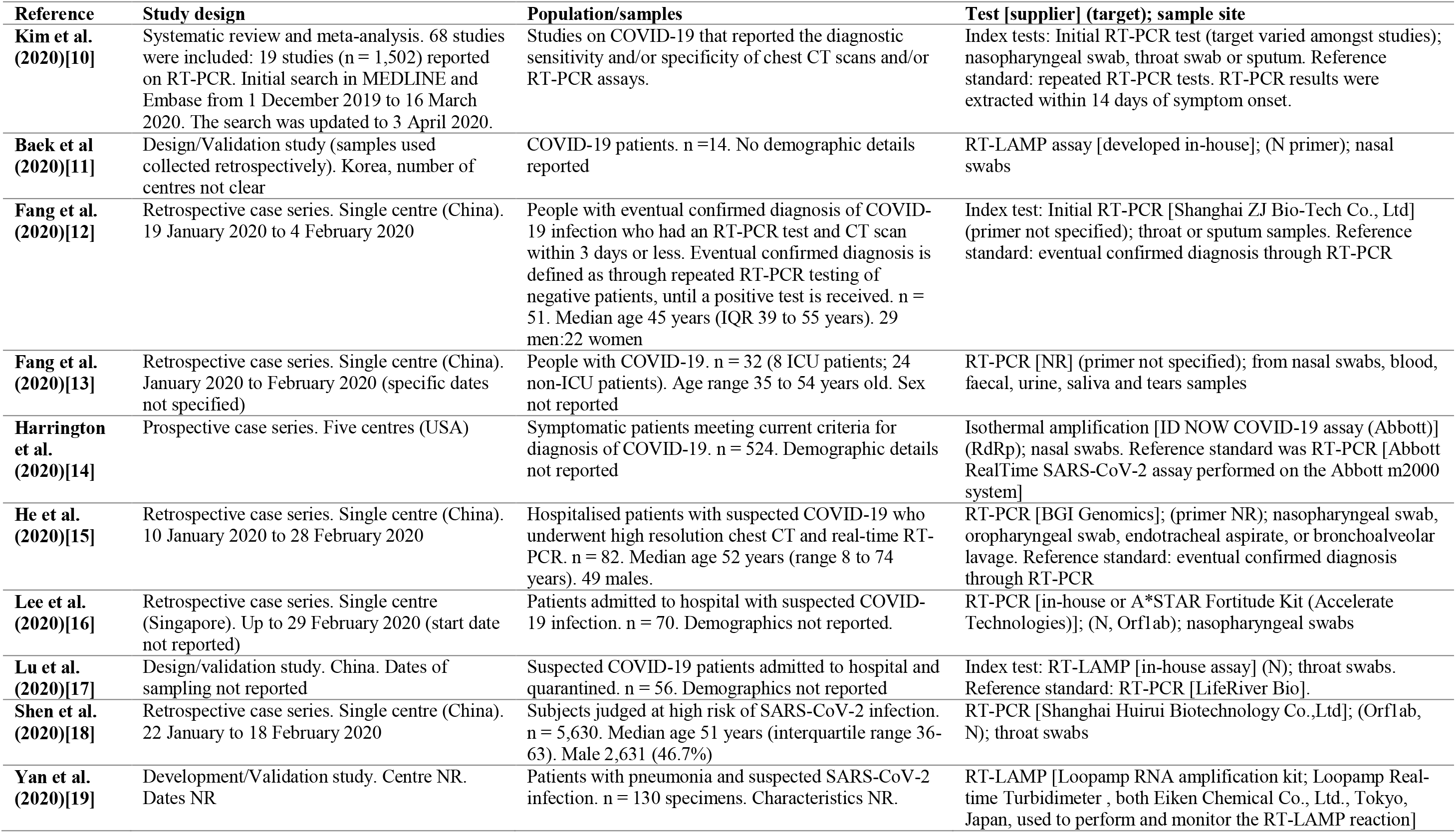
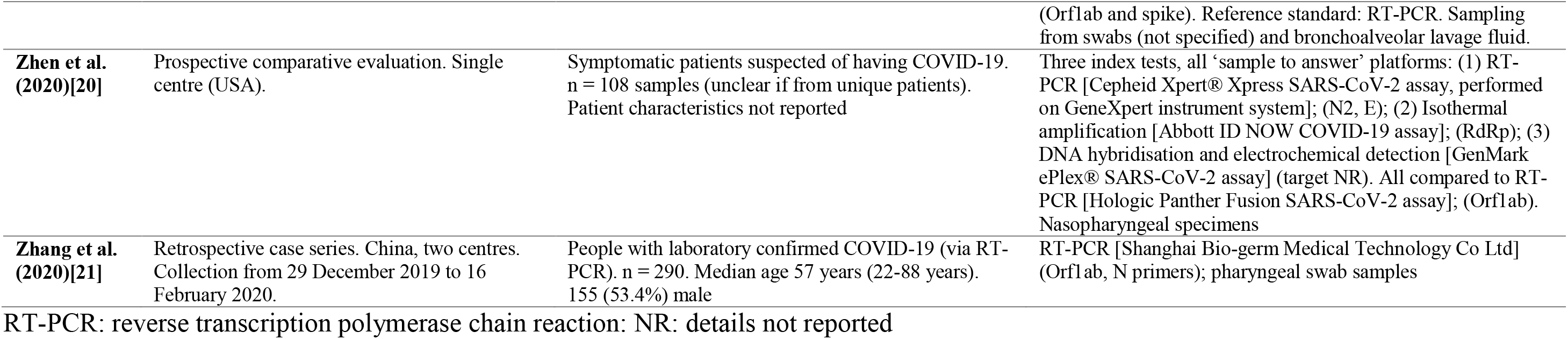
Characteristics of included studies reporting diagnostic accuracy outcomes for virus tests.

**Table 2.** Characteristics of included studies reporting diagnostic accuracy outcomes for antibody tests.

| Reference | Study design | Population/samples | Test [supplier] (target); sample site |
| --- | --- | --- | --- |
| **Cassaniti et al. (2020)[22]** | Cohort study. Single centre (Italy). Collection date NR. | 3 cohorts: (1) healthy volunteers with negative RT-PCR for COVID-19; (2) hospitalised patients with positive COVID-19 RT-PCR; (3) patients with suspected COVID-19 at their first access at emergency room. n = 110 (30 healthy volunteers; 30 COVID-19 patients; 50 patients with suspected COVID-19). Baseline characteristics reported separately for each cohort. | VivaDiag COVID-19 IgM/IgG Rapid point-of-care lateral flow immunoassay [Vivachek] (target NR); serum or blood samples. Serum samples were obtained at median 7 days (IQR 4 to 11 days) after positive result for hospitalised patients. Reference/Comparator: RT-PCR (RdRp and E primers); respiratory samples |
| **Dohla et al. (2020)[23]** | Single centre (Germany). Dates NR. | People within a community setting (high-prevalence area), presenting with COVID-19 symptoms (n = 39) and people diagnosed with COVID-19 (n = 10). Median age 46 year (IQR 28 to 72 years). 29/49 female (49.0%). | IgG/IgM point-of-care test [NR] (target NR); fingertip prick blood or serum. Reference standard: repeated RT-PCR [Altona diagnostics] (target NR); throat swabs. Serum samples for previously diagnosed individuals were also analysed using the antibody test. |
| **Hoffman et al. (2020)[24]** | Validation study. Centre NR. Study dates NR | Patients with confirmed COVID-19 or convalescents (n = 29). Controls: healthy volunteers without any known history of COVID-19 (n = 24); blood donor sera from health adults (n = 80) and babies (m = 20) collected during 2018 | IgG/IgM Rapid Test Cassette [Zhejiang Orient Gene Biotech Company] (target NR); blood/serum samples. Reference standard: RT-PCR |
| **Jin et al. (2020)[25]** | Retrospective study. Single centre (China) | People with a laboratory confirmed SARS-CoV-2 infection in hospital, and at least one viral serological test. (n = 43). Median age 47.0 years (IQR 34.0 to 59.0 years). 39.5% male. Control group: patients with suspected SARS-CoV-2 infection who were excluded and quarantined at home. (n = 33). Median age 31.0 years (IQR 25.5 to 37.5 years). 66.7% male. Suspected infected patients were discharged from hospital when they received two negative PCRs, performed in a 24 hour interval. | IgM and IgG chemiluminescence assay (CLIA) [Shenzhen YHLO Biotech Co., Ltd] (targets N protein and spike protein). Reference standard: confirmed diagnosis from RT-PCR (target not specified); sampling not clearly reported but includes oral swabs, anal swabs and sputum. Duration between first symptoms and serological test (CLIA) was 18 days (IQR 11 to 23 days) in the COVID-19 group, 3.0 days (2.0 to 8.0 days). |
| **Li et al. (2020)[26]** | Prospective development study. Single centre (China). 12 February 2020 to 20 February 2020 | People with suspected (RT-PCR negative) or confirmed (RT-PCR positive) COVID-19. n = 278 (89 confirmed; 189 probable). n = 273 controls were included. Baseline characteristics NR. | IgM and IgG colloidal gold assay [NR] (targets serum antibodies against N protein); serum specimens. RT-PCR assumed to be the reference standard (described as a 'control' by the authors); primer/target and sampling methods not known. |
| **Li et al. (2020)[27]** | Prospective development study. 8 centres (China). Dates NR | People with suspected COVID-19. n = 525 specimens (397 clinical positive; 128 clinical negative). Characteristics NR. | IgM/IgG rapid point-of-care lateral flow immunoassay [Jiangsu Medomics Medical Technologies] (targets antibodies against spike protein); blood (including serum and plasma). Reference standard: RT-PCR; respiratory specimens |
| **Liu et al. (2020)[28]** | Prospective study. Single centre (China). 18 January to 26 February 2020 | Hospitalised patients diagnosed with COVID-19. All patients were laboratory confirmed by RT-PCR. n = 314 (214 patients; 100 healthy controls). Baseline characteristics NR | IgM ELISA IgG ELISA [NR] (targets antibodies against N and spike); serum. Median time of sample collection was 15 days (range 0 to 55) |
| **Shen et al. (2020)[29]** | Prospective cohort study. Single centre | People with suspected COVID-19. Suspected COVID-19 was defined as a pneumonia that had related epidemiological history and fulfilled two of the following: fever and/or respiratory symptoms; imaging | IgM/IgG colloidal gold immunochromatography antibody kit [Shanghai Outdo Biotech Company] (target NR); blood. Reference standard: RT-PCR (target NR); nasopharyngeal and oropharyngeal swabs. At least 2 |
|  | (China). 20 January and 2 February 2020 | indicative of pneumonia; low/normal white cell count or low lymphocyte count. n = 150. Median age: PCR positive group 38 years (IQR 46 – 56 years); PCR negative group 32 years (IQR 20 to 42.5). Sex: PCR positive group 60.8% male; PCR negative group 56.6% male. Control: healthy donors (n = 26) | different samples were obtained from each patient for RT-PCR. If the result was inconclusive, repeated sample collection was required. A patient with at least 1 positive RT-PCR was confirmed as positive. Patients with two consecutive negative results were defined as PCR negative, but would only be diagnoses as non-COVID-19 if the symptoms could be explained by another condition or infection resolved following the corresponding treatments. Any other PCR negative result was treated as inconclusive. |
| **Spicuzza et al. (2020)[30]** |  | People with confirmed COVID-19 (n = 23) or suspected COVID-19 (n = 7). Confirmed COVID-19 was defined as consistent radiological/clinical findings, with positive RT-PCR. Suspected COVID-19 was defined as suggestive radiological/clinical findings but negative RT-PCR. Control: asymptomatic controls with negative RT-PCR (n = 7). n = 37. Mean age: confirmed COVID-19 57±17 years; suspected COVID-19 67±15 years. Controls age NR. Sex characteristics NR. | IgG/IgM POC Antibody Rapid Test Kit [Beijing Diagreat Biotechnologies Company] (spike); blood/serum/plasma. Reference standard: RT-PCR [NR] (target NR); nasopharyngeal swab or bronchial aspirate. |
| **Xiang et al. (2020)[31]** | Single centre (China). 19 January to 2 March 2020 | People with suspected (n = 24) or confirmed (n = 85) COVID-19. Diagnosis of laboratory confirmed COVID-19 was defined as positive nucleic acid tests for SARS-CoV-2 by RT-PCR. Diagnosis of suspected COVID-19 was based on negative RT-PCR, but satisfying 1 of the epidemiological history criteria and 2 of the clinical criteria. Control: samples from healthy blood donors or from hospitalised patients with other diseases. (n = 60) | IgM/IgG ELISA [Livzon Inc.] (NR); serum. Serum samples were obtained at different time periods after symptom onset. Reference standard: RT-PCR (ORF1ab and N); nasopharyngeal and/or oropharyngeal swabs |
| **Xu et al. (2020)[32]** | Retrospective study. Single centre (China). 20 January 2020 to 17 February 2020 | Patients with suspected COVID-19. n = 284 participants: 186 COVID-19 patients with RT-PCR positive result; 19 COVID-19 cases diagnosed by clinical symptoms; 79 controls with other diseases (negative RT-PCR). Baseline characteristics NR | IgM and IgG fully automated assay [NR] (target NR); serum samples. Comparator: RT-PCR. Reference standard: Diagnosis through positive RT-PCR or clinical symptoms. |
| **Zhao et al. (2020)[33]** | Retrospective study. Single centre (China). 11 January 2020 to 9 February 2020 | People with COVID-19. All enrolled cases were confirmed to be infected with SARS-CoV-2 by RT-PCR. n = 173 patients (535 samples). Median age 48 years (IQR 35 to 61). 51.4% female | Index tests: IgM ELISA [Beijing Wantai Biological Pharmacy Enterprise Co.,Ltd] (spike protein). IgG ELISA (N). Total antibody (Ab) ELISA (spike protein); plasma samples. Comparator: RT-PCR result. Reference standard: confirmed COVID-19 through positive RT-PCR |

CLIA: Chemiluminescent immunoassay; dpo: days post onset; ELISA: enzyme-linked immunosorbent assay; GICA: gold immunochromatography assay; IgA: Immunoglobulin A; IgG: Immunoglobulin G; IgM: Immunoglobulin M; LFIA: lateral flow immunoassay; RT-PCR: reverse transcription polymerase chain reaction; NR: details not reported

All the articles that reported on virus detection were based on the detection of amplified viral SARS-CoV-2 nucleic acid sequences. Most studies used laboratory-based RT-PCR tests conducted using standard in-house or commercially available reagents and equipment, although in some cases, assay details were not reported. The RT-PCR primer used (i.e. which part of the viral RNA is targeted and amplified) varied between studies, although again in some cases, primer details were not reported. In addition to RT-PCR, we identified five studies reporting the diagnostic performance of isothermal amplification assays.

The antibody tests studied used a range of different assay methods to detect one or more antibody type (different immunoglobulin classes and/or antibody targeted). In seven of the studies, tests were laboratory-based (enzyme-linked immunosorbent assay [ELISA]).[28,31,33-37] We identified 17 studies using assays (lateral-flow immunoassay [LFIA]; chemiluminescent immunoassay [CLIA]; colloidal gold immunochromatographic assay [GICA]) that could be suitable for point-of-care use,[22-27,29,30,34,38-45] but the tests were not applied at point-of-care, or it was not clearly reported that the test had been applied at point-of-care, in 14 of these studies. In two studies the type of assay was unclear.[32,46]

The reliability and applicability of each study’s conduct and reporting was assessed using the QUADAS-2 tool.[8] Virus tests and antibody tests were assessed separately and summary judgements are shown in Figure 2; signalling questions used and judgements per study are shown in Supplementary Appendix 3.

**Figure 2.**
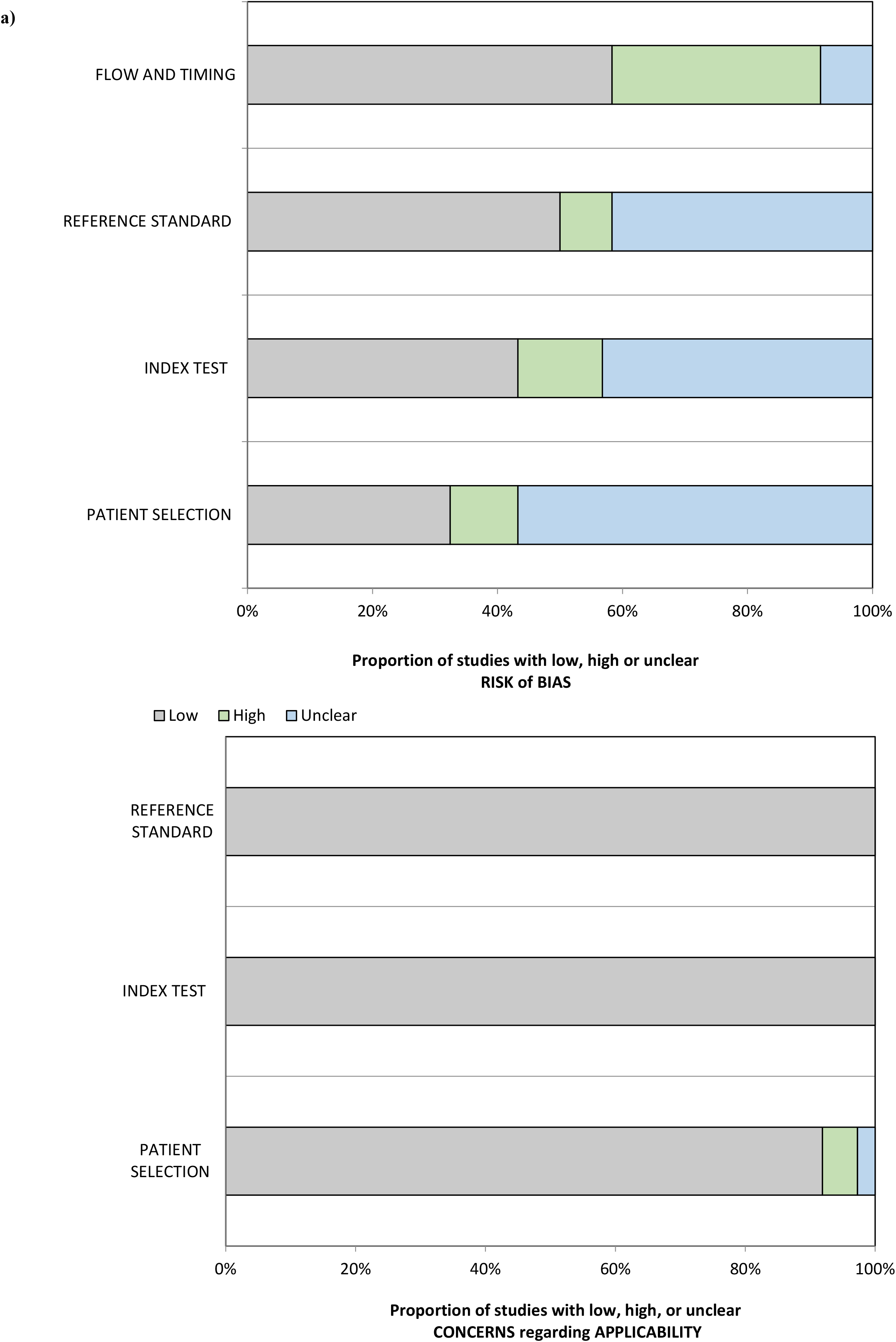
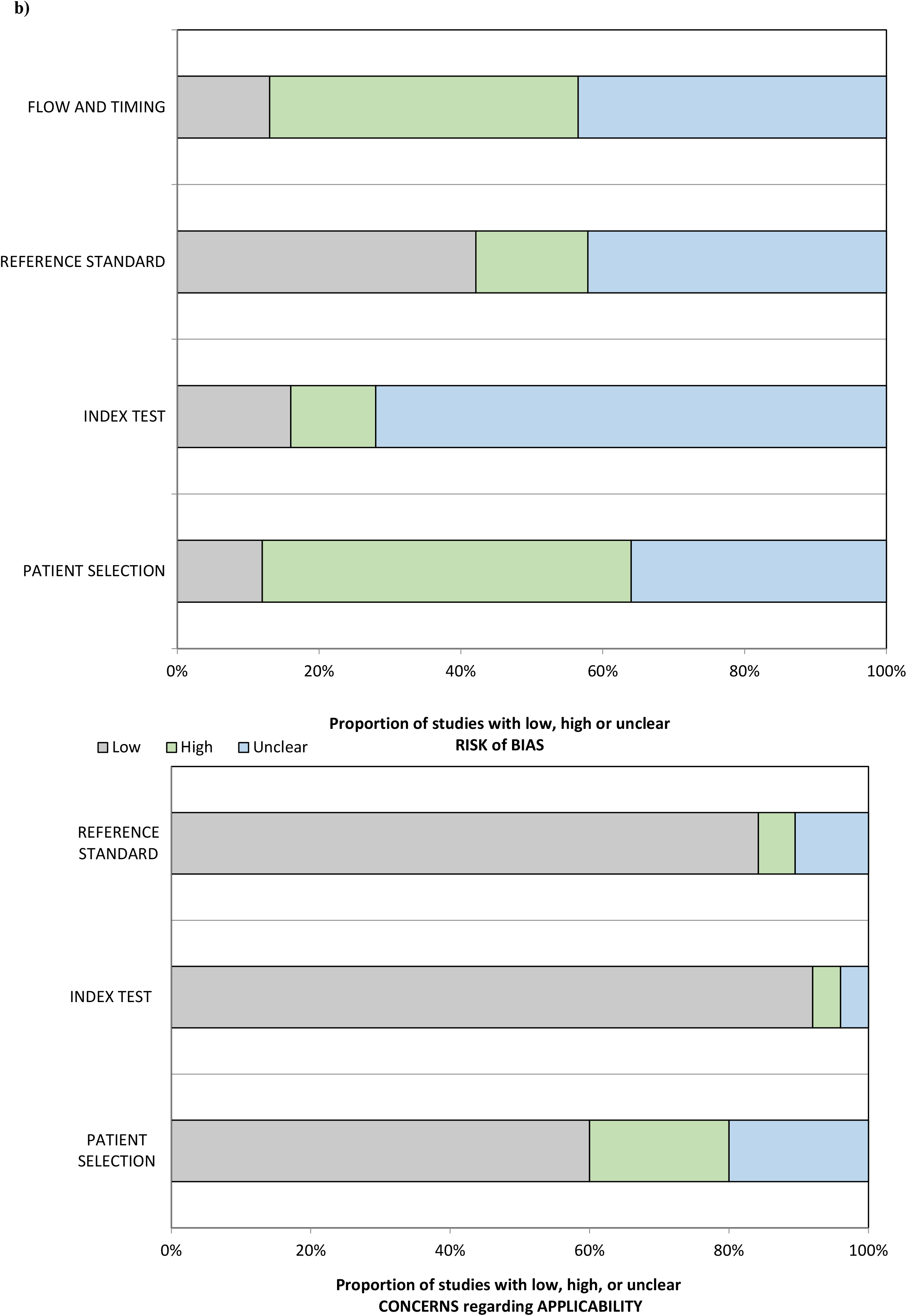
QUADAS2 risk of bias judgements. Summary of risk of bias and applicability assessments for (A) virus tests and (B) antibody tests.

For virus tests, most studies were judged to be of high or unclear risk of bias regarding patient selection, either because patients were selected for the study in a way that could have introduced bias (11% of studies) or because the method of patient selection was unclear (56% of studies). Risk of bias regarding how the index test was conducted or interpreted was judged to be high or unclear for 14% and 44% of studies, respectively, either because aspects of how the tests were conducted were unclear, or because tests were not conducted in a uniform manner. For the 12 studies that included a reference standard, we judged the risk of bias to be unclear in 42% and to be high in 8%, largely because not all tests were compared against a uniform reference standard, or some details of the reference standard were uncertain.

For antibody tests, the method of patient selection was judged to be unclear for 36% of studies and high for a further 52%. There was an unclear and high risk of bias regarding how the index test was conducted or interpreted in 72% and 12% of studies, respectively. For the 19 studies that included a reference standard, 42% were judged to have an unclear risk of bias and 16% a high risk of bias.

We identified one existing published meta-analysis estimating the sensitivity of an initial RT-PCR test, using the results of repeated RT-PCR tests as the reference standard.[10] This included studies published up to 3 April 2020, and aligned closely with our inclusion criteria regarding virus tests, although the authors included studies of any population size, whereas we elected to exclude studies that included less than ten patients, meaning we omitted seven studies (total 46 patients) that were included in the earlier meta-analysis. We used data from this analysis and studies published subsequently to determine that the overall sensitivity of RT-PCR is 87.8% (95% confidence interval 81.5% to 92.2%), based on 16 studies of 3818 patients. Because these studies only included cases where COVID-19 was confirmed to be present, specificity cannot be legitimately estimated.

Five studies (972 patients or samples in total) reported the diagnostic accuracy of isothermal amplification assays in the diagnosis of patients with suspected COVID-19, using test results from RT-PCR as a reference standard.[11,14,17,19,20] Because the use of a single RT-PCR test as a reference standard may not be representative of true disease presence, we deemed it inappropriate to use the results of these studies to derive a single pooled estimate of sensitivity and specificity. Reported diagnostic sensitivity and specificity estimates range from 74.7% to 100% and 87.7% to 100%, respectively. Table 3 provides a detailed breakdown of results.

**Table 3.** Diagnostic accuracy outcomes for virus tests.

| Reference | Assay and target | Number of patients/samples | Index test; Comparator (if applicable) |
| --- | --- | --- | --- |
| *Pooled sensitivity* |  |  |  |
| **Updated analysis including results from Kim et al. (2020)[10] systematic review and meta-analysis and subsequent publications.** | NR | n = 3,818; 16 studies | RT-PCR: 87.8% (95% CI 81.5%, 92.2%); |
| *Sensitivity for studies that could not be pooled* |  |  |  |
| **Baek et al. (2020)[11]** | N | n = 154 samples | RT-LAMP: 100% |
| **Harrington et al. (2020)[14]** | RdRp | n = 524 patients | Isothermal amplification (Abbott ID-NOW assay): 74.7% (95% CI 67.8% to 80.8%) |
| **Lu et al. (2020)[17]** | N | n = 56 patients | RT-LAMP: 94.4% (95% CI 81.3 to 99.3%) |
| **Yan et al. (2020)[19]** | Orflab and spike | n = 130 samples | RT-LAMP: 100% (95% CI 92.3% to 100%) |
| **Zhen et al. (2020)[20]** | N2, E | n = 108 samples | RT-PCR [Cepheid Xpert® Xpress SARS-CoV-2 assay: 98.3% (95% CI 90.7% to 99.9%) |
|  | RdRp | n = 108 samples | Isothermal amplification [Abbott ID NOW COVID-19 assay]: 87.7% (95% CI 76.3% to 94.9%) |
|  | NR | n = 108 samples | DNA hybridisation and electrochemical detection [GenMark ePlex®: 98.3% (95% CI 90.7% to 99.9%) |
| *Specificity* |  |  |  |
| **Baek et al. (2020)[11]** | N | n = 154 samples | RT-LAMP: 98.7% |
| **Lu et al. (2020)[17]** | N | n = 56 patients | RT-LAMP: 90.0% (95% CI 68.3% to 98.8%) |
| **Harrington et al. (2020)[14]** | RdRp | n = 524 patients | Isothermal amplification (Abbott ID-NOW assay): 99.4% (95% CI 97.8% to 99.9%) |
| **Yan et al. (2020)[19]** | Orflab and spike | n = 130 specimens | RT-LAMP: 100% (95% CI 93.7% to 100%) |
| **Zhen et al. (2020)[20]** | N2, E | n = 108 samples | RT-PCR [Cepheid Xpert® Xpress SARS-CoV-2 assay: 98.3% (95% CI 92.3% to 100%) |
|  | RdRp | n = 108 samples | Isothermal amplification [Abbott ID NOW COVID-19 assay]: 87.7% (95% CI 92.3% to 100%) |
|  | NR | n = 108 samples | DNA hybridisation and electrochemical detection [GenMark ePlex®: 100% (95% CI 92.3% to 100%) |

CI: confidence interval; RT-LAMP: reverse transcription loop-mediated isothermal amplification; RT-PCR: reverse transcription polymerase chain reaction; NR: details not reported.

Ten studies on antibody testing (757 participants included; number not clear for two studies) reported sensitivity and specificity[22,23,25-27,29-33] or sufficient information to allow these to be calculated. Two additional studies reported specificity only.[24,28] Where a reference standard was included, this was usually RT-PCR (initial and repeats until a positive confirmation); one study that used either RT-PCR or clinical diagnosis to determine final disease status. Furthermore, studies used a range of different antibody types and targets. Because of these limitations, we concluded that pooling data across studies was not appropriate.

The reported sensitivity in these studies ranged from 18.4% to 96.1%. Notably, the lowest reported sensitivity was using a point-of-care test,[22] although sensitivity figures below 50% were also reported for one laboratory test.[25] Specificity was reported in 12 studies (682 participants included; number not clear for two studies) and ranged from 88.9% to 100%. Full outcomes from these studies are shown in Table 4.

The positive predictive value (PPV) and negative predictive value (NPV) of RT-PCR was estimated at different prevalence levels. We used our pooled sensitivity estimate of 87.8%, and because we were unable to calculate specificity using the evidence found, we used a previously published estimate of 98.0% for specificity[47]. Prevalence estimates were based on data from Public Health England (PHE).[48] A prevalence rate of 3.0% was estimated based on PHE data up to 6^th^ August 2020, which showed that there have been 308,134 confirmed cases of COVID-19 from 10,236,970 tests. At this prevalence level, RT-PCR testing was estimated to have a PPV of 57.7% and NPV of 99.6%. To estimate the utility of the test at times of high prevalence, PPV and NPV were also estimated using PHE data up to 1^st^ May 2020 (the date at which the daily number of cases was at its highest point). On this date, a prevalence rate of 24.6% was estimated based on 177,454 confirmed cases of COVID-19 from 721,124 tests. At this prevalence level, RT-PCR testing was estimated to have a PPV of 93.5% and NPV of 96.1%.

**Table 4.** Diagnostic accuracy outcomes for antibody tests.

| Reference | Assay and target | Number of patients/samples | Index test; Comparator (if applicable) |
| --- | --- | --- | --- |
| *Sensitivity* |  |  |  |
| Cassaniti et al. (2020)[22] | LFIA , VivaChek POC | n = 50 patients (suspected cases only) | IgM/IgG: 18.4% |
| Dohla et al. (2020)[23] | IgM/IgG POC test | n = 49 | IgM/IgG: 36.4% (95%CI 17.2; 59.3) |
| Jin et al. (2020)[25] | CLIA (N and spike proteins) | n = 27 | IgM: 48.1% (13/27)<br>IgG: 88.9% (24/27) |
| Li et al. (2020)[26] | colloidal gold | Population not clear | IgM: 78.7%;<br>IgG: 73.0%<br>IgM/IgG: 87.6% |
| Li et al. (2020)[27] | LFIA, Jiangsu Medomics POC | n = 525 specimens | IgM/IgG: 88.66% |
| Spicuzza et al. (2020)[30] | LFIA (spike) | n = 37 | IgG/IgM: 82.6% |
| Shen et al. (2020)[29] | Colloidal gold (NR) | n = 150 | IgM/IgG: 71.1% [95% CI 0.609-0.797] |
| Xiang et al. (2020)[31] | ELISA (NR) | n = 66 | IgM: 77.3% (51/66)<br>IgG: 83.3% (55/66) |
| Xu et al. (2020)[32] | Fully-automated assay (NR) | n = 205 patients | IgM: 70.24%(144/205)<br>IgG: 96.10%(197/205) |
| Zhao et al. (2020)[33] | ELISA (spike for IgM and Ab; N for IgG) | n = 173 samples | IgM: 82.7% (143/173);<br>IgG: 64.7% (112/173);<br>Ab: 93.1% (161/173);<br>RT-PCR: 67.1%* (112/?) |
| *Specificity* |  |  |  |
| Cassaniti et al. (2020)[22] | LFIA , VivaChek POC | n = 50 (suspected cases only) | IgM/IgG: 91.7% |
| Li et al. (2020)[26] | colloidal gold | Population not clear | IgM: 98.2%;<br>IgG: 99.3%;<br>IgM/IgG: 98.2% |
| Li et al. (2020)[27] | LFIA, Jiangsu Medomics POC | n = 525 specimens | IgM/IgG: 90.63%. |
| Liu et al. (2020)[28] | ELISA (spike) | n = 100 healthy controls | IgM: 100% (0/100);<br>IgG: 100% (0/100)<br>IgM and/or IgG: 100% (0/100) |
| Xu et al. (2020)[32] | Fully-automated assay (NR) | n = 79 patients | IgM: 96.20% (76/79)<br>IgG: 92.41%(73/79) |
| **Zhao et al. (2020)[33]** | ELISA (spike for IgM and Ab; N for IgG) | Not clear | Total Ab: 99.1% (211/213);<br>IgM: 98.6% (210/213);<br>IgA: 99.0% (195/197) |
| **Jin et al. (2020)[25]** | CLIA (N and spike proteins) | n = 33 | IgM: 100% (33/33)<br>IgG: 90.9% (30/33) |
| **Xiang et al. (2020)[31]** | ELISA (NR) | n = 60 | IgM: 100% (60/60);<br>IgG: 95.0% (57/60) |
| **Dohla et al. (2020)[23]** | IgM/IgG POC test | n = 49 | IgM/IgG: 88.9% (95% CI 70.8; 97.7) |
| **Spicuzza et al. (2020)[30]** | LFIA (spike) | n = 37 | IgG/IgM 92.9% |
| **Hoffman et al. (2020)[24]** | LFIA (NR) | n = 124 (controls) | IgM: 100% (0/124);<br>IgG: 99.2% (1/124) |
| **Shen et al. (2020)[29]** | Colloidal gold (NR) | n = 150 | IgM/IgG: 96.2% [95% CI 0.859-0.993] |

CI: confidence interval; CLIA: Chemiluminescent immunoassay; ELISA: enzyme-linked immunosorbent assay; GICA: gold immunochromatography assay; IgA: Immunoglobulin A; IgG: Immunoglobulin G; IgM: Immunoglobulin M; LFIA: lateral flow immunoassay; RT-PCR: reverse transcription polymerase chain reaction; NR: details not reported.

We identified 18 studies[13,43,49-64] that compared RT-PCR for SARS-CoV-2 results from samples taken from different parts of the body. These results are presented in Table 5. Most samples were taken from the upper respiratory tract (Supplementary Appendix 4 summarises detection rates for individual sites in the upper respiratory tract, where reported). Other sample sites were saliva, sputum and stool/rectal swab. The detection rates varied across sample sites but the heterogeneous nature of the studies makes meaningful comparison difficult. Detection rates were consistently low with urine or tears/conjunctiva sampling. Detection rates in blood samples were mixed, with some studies reporting very low detection rates, whilst others reported rates that were comparable to samples from other sites in the same population.

**Table 5.**
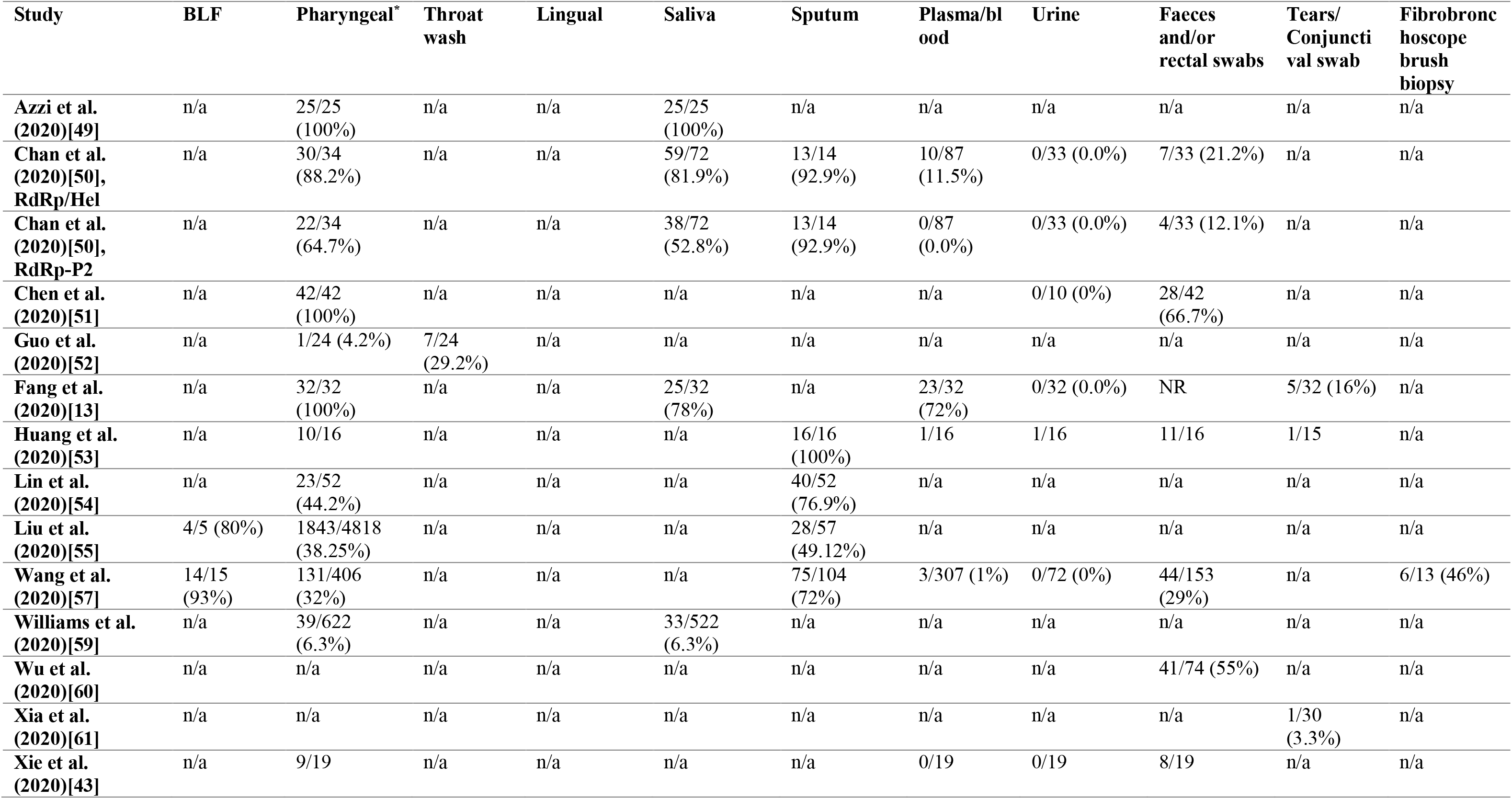
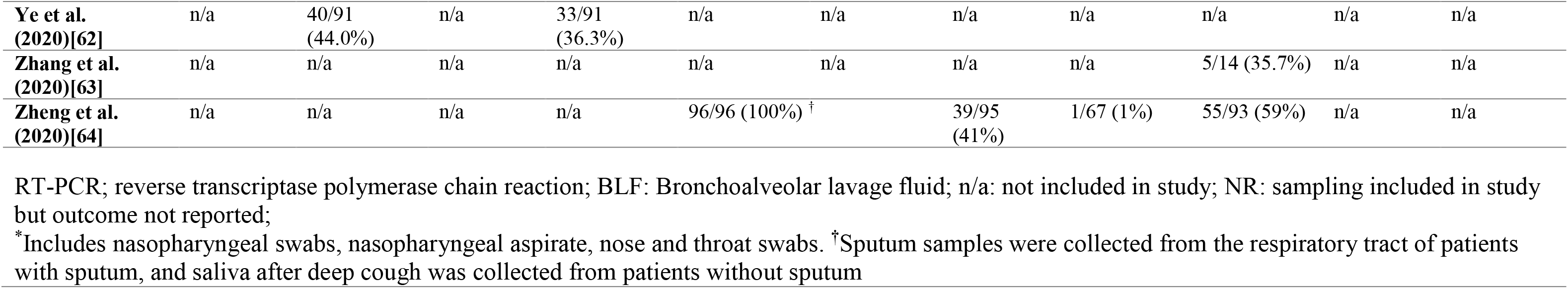
Virus test detection rates in studies comparing different sample sites.

The majority of studies tested people with relatively severe disease and a high suspicion of COVID-19 infection. Considering other populations, two studies[65,66] tested UK healthcare workers and three studies[18,67,68] tested people outside of hospital (or as outpatients). One further study[69] routinely tested pregnant women. All these studies only reported detection rates: results are summarised in Supplementary Appendix 4.

Ten studies provided data on antibody detection (seroprevalence) at different points in time after the onset of confirmed COVID-19 disease.[28,33-36,39-41,44,46] Detailed results are shown in Supplementary Appendix 4.

## DISCUSSION

This review summarises the available published evidence of the effectiveness of tests that are used in the diagnosis of current or previous COVID-19 infection up to 4 May 2020. Despite this work taking place relatively early in the COVID-19 pandemic, 38 published studies were identified that reported on the effectiveness of tests for detecting the presence of SARS CoV-2 virus and 25 studies were identified that reported on testing for the presence of antibodies. Analysis of these studies using the QUADAS-2 framework revealed high or unclear risks of bias in the majority, most commonly as a result of unclear methods of patient selection and test conduct, or because of the use of a reference standard that may not definitively diagnose COVID-19. Nonetheless, the available evidence provides information on which to begin to judge the possible clinical effectiveness of COVID-19 testing, although significant uncertainties remain in the evidence base regarding their clinical and public health application.

In the course of our work, the first meta-analysis of diagnostic accuracy for SARS-CoV-2 virus tests was published by Kim et al.[10] This included pooled analysis of 19 studies (1,502 patients) and used the results of repeated laboratory-based RT-PCR as the reference standard. This aligned closely with our own inclusion criteria for virus tests, although Kim et al. included studies of any population size, we excluded studies with less than ten patients, meaning we omitted seven studies (46 patients). However, by including more recently published studies in our analysis the number of patients more than doubles from 1,502 to 3,818 patients. The analysis by Kim et al. estimated that the sensitivity of an initial RT-PCR test is 89% (95% CI 81% to 94%). Our addition of data from more recent studies leads us to conclude a sensitivity of 87.8% (95% CI 81.5% to 92.2%).An analysis estimating the PPV and NPV of RT-PCR showed that the NPV is likely to be high while PPV may be low at times where the prevalence in the tested population is low. The likely prevalence in the tested population should therefore be a key consideration for decision makers when interpreting test results and deciding on testing strategies. Despite our finding of a high NPV for RT-PCR, uncertainty may remain with a negative test result, especially in the context of high clinical suspicion, and the possibility of a false negative result also needs to be considered. Possible causes for false negative tests include laboratory error, sampling error, and variability in viral shedding with the lack or negligible presence of virus nucleic acid in the tissue sampled at the time of sampling. Determining the specificity of SARS-CoV-2 nucleic acid testing is particularly challenging because of the inclusion in the published studies of patients considered to be suffering from COVID-19 as well as the lack of a reference standard that validates the absence of disease. The assessment of overall diagnostic accuracy in laboratory testing for the presence of the SARS-CoV-2 virus is hampered by the absence of a definitive reference standard and by a wide range of target primers, methods and types of sampling used in the published studies. Additionally, there is very limited published information on the diagnostic accuracy of point-of-care or near-patient tests.

Of the 25 studies that assessed antibody tests, 10 reported diagnostic accuracy in terms of both sensitivity and specificity, almost all using RT-PCR (initial or repeat testing) as the reference standard.[18,22,23,25-27,30-33] Accepting the limitations already discussed around the absence of a diagnostic reference standard, the overall sensitivity reported in these studies varied widely, from 18.4% to 96.1% although the specificity was more consistent and ranged from 88.9% to 100%. The clinical implications of these data are that considerable uncertainty remains about the implications of a negative antibody test with a significant possibility of false negativity, while the presence of a positive antibody test carries with it a high likelihood of previous COVID-19 infection. There is very limited information available on the accuracy of point-of-care antibody tests.

Our study has some limitations, primarily due to the nature of the evidence found by our searches. The rapid nature of this work (to help inform decision makers at the outset of the COVID-19 pandemic in the United Kingdom) meant some steps in a full systematic review were not completed: there was minimal consultation with decision makers on the inclusion and exclusion criteria for the review, and we did not publish our protocol in advance of commencing the review. Other limitations relate to the nature of the evidence we found, and that this work was completed during the early stages of the COVID-19 pandemic. The lack of a recognised reference standard meant we considered studies for inclusion that used any appropriate method to verify test results. Whilst initial suspicion of COVID-19 may be based on clinical assessment combined with radiological results, WHO advice is that laboratory-based nucleic acid testing (such as RT-PCR) should be used to confirm cases with further confirmation by nucleic acid sequencing when necessary or feasible.[70] The only suitable studies that allowed the diagnostic accuracy of RT-PCR to be assessed compared initial test results to repeated RT-PCR testing in the same individuals: this allowed us to estimate the sensitivity of an initial RT-PCR test, using final (positive) results of the repeated test as the reference standard. Use of this reference standard, which only validates the presence of disease and not its absence, means specificity cannot be determined. We estimated the PPV and NPV of RT-PCR and at different prevalence rates; estimates from PHE were judged to provide the best current evidence for prevalence. However, it should be noted that there are limitations with this approach. Most notably, it is based upon the total number of tests rather than the number of people tested. As such, the estimates may underestimate prevalence as many people will have been tested more than once. Crucially, because we could not calculate specificity from the evidence found by our own systematic review, we relied on a previously published estimate of 98.0%. PPV is highly sensitive to this estimate, emphasising the need for further reliable published estimates of the sensitivity of RT-PCR to the interpretation of this test, particularly in low prevalence populations. Furthermore, the evidence included in this pooled analysis and other individual studies we identified used a range of target primers, methods and type of sampling.

We observed similar limitations with evidence on other tests. We found studies reporting diagnostic accuracy of antibody tests and of LAMP as a method of virus detection. However, the reference standard used was RT-PCR (initial and repeat tests), except for one study that used either RT-PCR or clinical diagnosis to determine final disease status. As already concluded, a true assessment of the accuracy of RT-PCR test results is very challenging, and using RT-PCR for validation means the same limitations apply to the results of any antibody or LAMP tests studied in this way. These tests also varied considerably in their conduct and protocols used. These limitations led us to conclude that it was inappropriate to conduct pooled analysis of diagnostic accuracy, meaning limited conclusions about antibody and LAMP tests can be drawn based on the data currently available. Lastly, for all types of test, there are a wide range of different commercially available testing products and kits, as well as some that use protocols developed in-house by academic and public health testing laboratories. Where available, we have detailed the exact test used for each data source (Table 1, Table 2, and Supplementary Appendix 2), but our evidence synthesis does not take into account similarities or differences between specific test kits or protocols, and the results should be interpreted with this in mind.

Alternative approaches to validating COVID-19 test results could use genomic sequencing, testing for multiple primer targets, confirmatory testing for other respiratory viruses, longterm follow-up, or clinical signs and symptoms. Each would have potential advantages and disadvantages. For example, genomic sequencing would determine the exact strain of virus present and could detect cases of infection where primer target regions were not conserved, resulting in a false negative RT-PCR result. However, sequencing could only be used to ‘rule in’ the presence of the virus and not to rule it out. Furthermore, it is time and resource intensive and so would be highly unlikely to be undertaken for all samples in routine practice. Future studies might need to use a combination of these factors as a composite reference standard that could validate results from both positive and negative diagnoses, allowing sensitivity and specificity of the test to estimated with greater certainty.

In applying the results of the published studies on testing for COVID-19 to influencing the development of evidence-based testing strategies, it should be noted that the majority of published studies reported on the results of COVID-19 virus or antibody testing were done in a hospital setting and in symptomatic patients with confirmed or suspected COVID-19 infection. Data on testing in other settings is comparatively limited. Only three studies[18,67,68] were identified that used RT-PCR to detect SARS-CoV-2 in the general population in the context of mild influenza-like symptoms while only two studies were found[65,66] that reported on the testing of UK healthcare workers. Furthermore, only one study[23] was identified that reported on the results of antibody testing outside of a hospital setting. In a rapidly developing pandemic, the widespread use of testing is an essential element in the development of effective public health strategies, but it is important to acknowledge the gaps that exist in the current evidence base and that, where possible, these should be addressed in future studies.

In regard to future research, more data is required to substantiate the effectiveness of tests to detect the presence of SARS-CoV-2 virus or antibodies to SAS-CoV-2 in different populations and more evidence is needed to compare the effectiveness of laboratory based testing and point-of-care testing strategies. Further clarity is required about the optimal timing of tests relative to symptom onset. The results of testing in people with no or minimal symptoms in a community-based setting needs further analysis and the impact of these data on public health measures needs to be fully analysed. Evidence should be prospectively collected during the implementation of public health strategies that combine testing with tracing and isolating individuals who have been in contact with COVID-19 sufferers. The UK National Institute for Health and Care Excellence (NICE) have recently published an evidence standards framework which describes a 3-stage approach to collecting the best possible data and evidence in the short and long term which is applicable both to established and developing COVID-19 tests.[71] The framework assumes that the tests’ analytical performance is already established, that developers are complying with existing quality systems for manufacturers (ISO 13485) and laboratories (ISO 15198 or 17025), and recommends that a good diagnostic accuracy study should be followed by demonstrating the clinical significance as well as the economic impact of applying the index test.

## Data Availability

n/a

## CONTRIBUTORS

DJ, MP, SM and PG conceived the project and prepared the review inclusion and exclusion criteria. JW prepared and ran search strategies with input from DJ. DJ and LE screened evidence, extracted data from relevant studies and carried out risk of bias assessments; data was independently checked by KC. DJ and MP analysed data to generate outcomes. DJ, LE, JW and PG wrote the manuscript with input and editing from all other authors.

## ACKNOWLEDGEMENTS

None

